# Feasibility of Bilateral Auricular Vagus Nerve Stimulation Combined with Exercise for Improving Cardiovascular and Motor Outcomes in Parkinson’s Disease

**DOI:** 10.1101/2025.08.06.25332986

**Authors:** Alexandra Evancho, Jennifer Dawson, Harrison Walker, Christopher Ballmann, William J. Tyler

## Abstract

**Background and Purpose:** Neuromodulation and physical therapy (PT) can both mitigate motor and non-motor symptoms of Parkinson’s Disease (PD). There are a lack of studies examining the integration of transcutaneous auricular vagus nerve stimulation (taVNS) with PT or exercise for improving Parkinsonian symptoms. The study was designed to investigate the safety, tolerability, and feasibility of combining bilateral taVNS with PT to enhance the therapeutic benefits of exercise as medicine in a clinical setting.

**Methods:** This pilot study was a randomized, sham-controlled clinical trial. Participants were randomly assigned to receive active or sham bilateral taVNS in combination with physical therapy for 12 visits over 6 weeks. We quantified safety, tolerability, and feasibility outcomes, and explored changes in cardiovascular and motor function over time.

**Results:** We observed taVNS was well tolerated without reported adverse events. We observed taVNS administered prior to physical therapy significantly decreased heart rate and blood pressure at rest. We observed the active taVNS treatment group exhibited more sustained improvements in motor function and balance compared to baseline. Due to the small size of the feasibility study, we did not detect between-group differences.

**Discussion and Conclusions:** Combining taVNS with physical therapy was safe, feasible, and well-tolerated. Preliminary results suggest taVNS has the potential to enhance rehabilitation outcomes by modulating cardiovascular function prior to and during PT and exercise. These findings support the need for larger clinical trials and real-world studies investigating the integration of taVNS into PT and exercise methods for improving PD symptomology.

**Trial Registration:** ClinicalTrials.gov NCT05871151

## 1. INTRODUCTION

Parkinson’s disease (PD) is a progressive neurodegenerative disease characterized by motor and non-motor symptoms [1–3]. Exercise is a well-established and effective non-pharmacologic therapy that reduces symptom severity and potentially attenuates disease progression [4, 5]. Physical Therapy (PT) is frequently prescribed for individuals with PD and provides a structured, reimbursable setting for exercise. However, translating evidence-based exercise protocols into routine PT care is often hindered by practical limitations and disease-specific barriers that are typically not accounted for during clinical trials [6, 7]. Several neuromodulation approaches have been combined with physical rehabilitation methods to enhance recovery and restore function for patients with a broad range of neurologic conditions and injury [8–10]. However, neuromodulation has not been systematically tested as a strategy to overcome PD-specific exercise barriers within standard PT practice.

Vagus nerve stimulation (VNS) is a neuromodulation method that may specifically enhance the efficacy of PT for individuals with PD. VNS is traditionally delivered via a surgically implanted pulse generator targeting the left cervical vagus nerve, however clinical translation of this approach is limited by stimulation-associated adverse events and poor scalability for routine rehabilitation. Transcutaneous auricular vagus nerve stimulation (taVNS) is a safe, non-invasive alternative to VNS that that can produce therapeutic outcomes by targeting the auricular branch of the Vagus Nerve (ABVN) through the skin of the external ear using low intensity, pulsed electrical currents [11–13]. Given the widespread use of associated transcutaneous electrical nerve stimulation (TENS) methods in PT, taVNS offers a practical, familiar, low-risk, and potentially scalable adjunct to rehabilitation interventions in PD [9].

The vagus nerve (CN X) is a cranial nerve that plays a major role in regulating autonomic nervous system (ANS) function including cardiovascular (CV) and immune function [14–18]. In PD, ANS dysfunction (dysautonomia) is highly prevalent, and can blunt the CV system’s response to exercise [19–22], which may limit the ability to achieve target heart rates during training and reduce the effectiveness of exercise interventions. Several recent studies in healthy volunteers demonstrate that taVNS can improve CV flexibility and performance in response to exercise [23, 24] and environmental stressors [25, 26]. Furthermore, cardiac vagal activity plays a causally deterministic role in our ability to exercise and achieve CV responses required to support physical and mental exertion [17, 27, 28]. Therefore, we hypothesize taVNS may acutely enhance CV dynamics to improve exercise capacity and tolerance in patients with PD. Additionally, the ABVN provides direct afferent pathways to the nucleus tractus solitarius (NTS), the locus coeruleus (LC), dorsal motor nucleus of the vagus, and other brainstem nuclei involved in autonomic regulation of psychophysiological arousal and neurophysiological tone [29, 30]. Activation of noradrenergic (NA) signaling pathways by the LC–norepinephrine (LC-NE) system modulates psychophysiological arousal, facilitates cortical plasticity, enhances attention, and improves motor learning [31–34]. It is also important to note, patients with PD often suffer progressive degeneration of NA signaling which can exacerbate symptoms including motor and cognitive dysfunction, as well as disrupt plasticity [1, 3, 35, 36]. Using taVNS to regulate CV dynamics and to modulate ascending NA signaling just prior to physical therapy may therefore “prime” the body for exercise by optimizing physiological adaptability during exertion while enhancing motor plasticity. This dual mechanism forms the basis for our hypothesis that taVNS can act synergistically with PT to improve motor outcomes in PD.

Studies investigating taVNS as a standalone therapy have demonstrated that it holds promise for alleviating motor and psychological symptoms associated with PD [37–41]. Additionally, several pioneering studies have shown taVNS used in conjunction with PT approaches can improve motor outcomes and somatic symptoms in other patient populations [42–47]. However, taVNS has not been investigated in conjunction with PT in individuals with PD. This pilot study aimed to establish and test a protocol for combining taVNS with PT in individuals with PD, with the primary goal of evaluating safety, tolerability, and feasibility. Our observations are intended to inform future trials investigating the potential therapeutic benefits of combining taVNS with PT and exercise as an intervention.

## 2. METHODS

### 2.1 Trial Design and Regulatory Oversight

This study was designed as a pilot, randomized, sham-controlled clinical trial with aims focused on assessing safety, tolerability, and feasibility. Blinding procedures were implemented to evaluate the feasibility of maintaining participant and assessor blinding for a future definitive trial and to reduce the risk of bias in exploratory outcome measures. The study protocol included 16 visits: screening, baseline motor assessment, 12 treatment sessions over six weeks, a post-test assessment, and a four-week follow-up visit (**Figures 1** and **2A**). All assessment visits were performed in the OFF-medication state (≥12 hours since last dopaminergic medication) to reduce the confounding influence of medication on motor scores. In contrast, all treatment sessions (including taVNS and PT) were completed in the ON medication state to reflect real-world clinical practice and ensure safety during exercise. Cognitive and quality of life outcomes were collected at each assessment but are beyond the scope of this paper. Participants were randomized at a 1:1 allocation ratio into 1 of 2 groups: (1) active taVNS plus PT or (2) sham taVNS plus PT. Written informed consent was obtained prior to enrollment and study participation. The study protocol was approved by the University of Alabama at Birmingham IRB and registered on ClinicalTrials.gov (NCT05871151).

**Figure 1.**
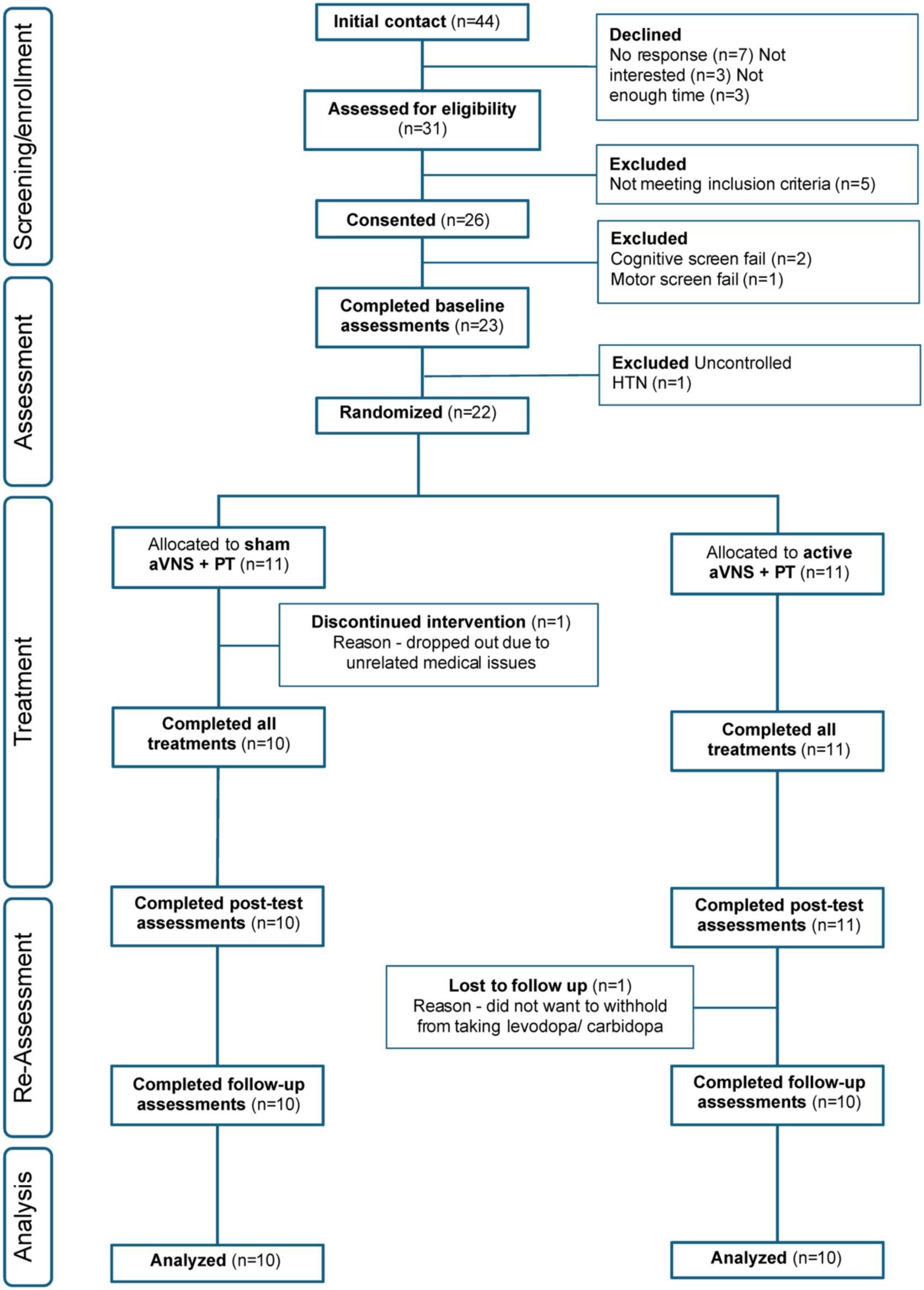
CONSORT Flow Diagram. The CONSORT flow diagram for the study illustrates participant screening, enrollment, randomization, intervention, and analysis. Of 44 individuals initially contacted, 22 were randomized into either sham or active transcutaneous auricular vagus nerve stimulation (taVNS) treatment groups. A total of 20 participants completed all study procedures including 12 taVNS plus physical therapy treatment sessions and were included in the final analysis, with reasons for exclusions and attrition documented throughout the flow.

**Figure 2.**
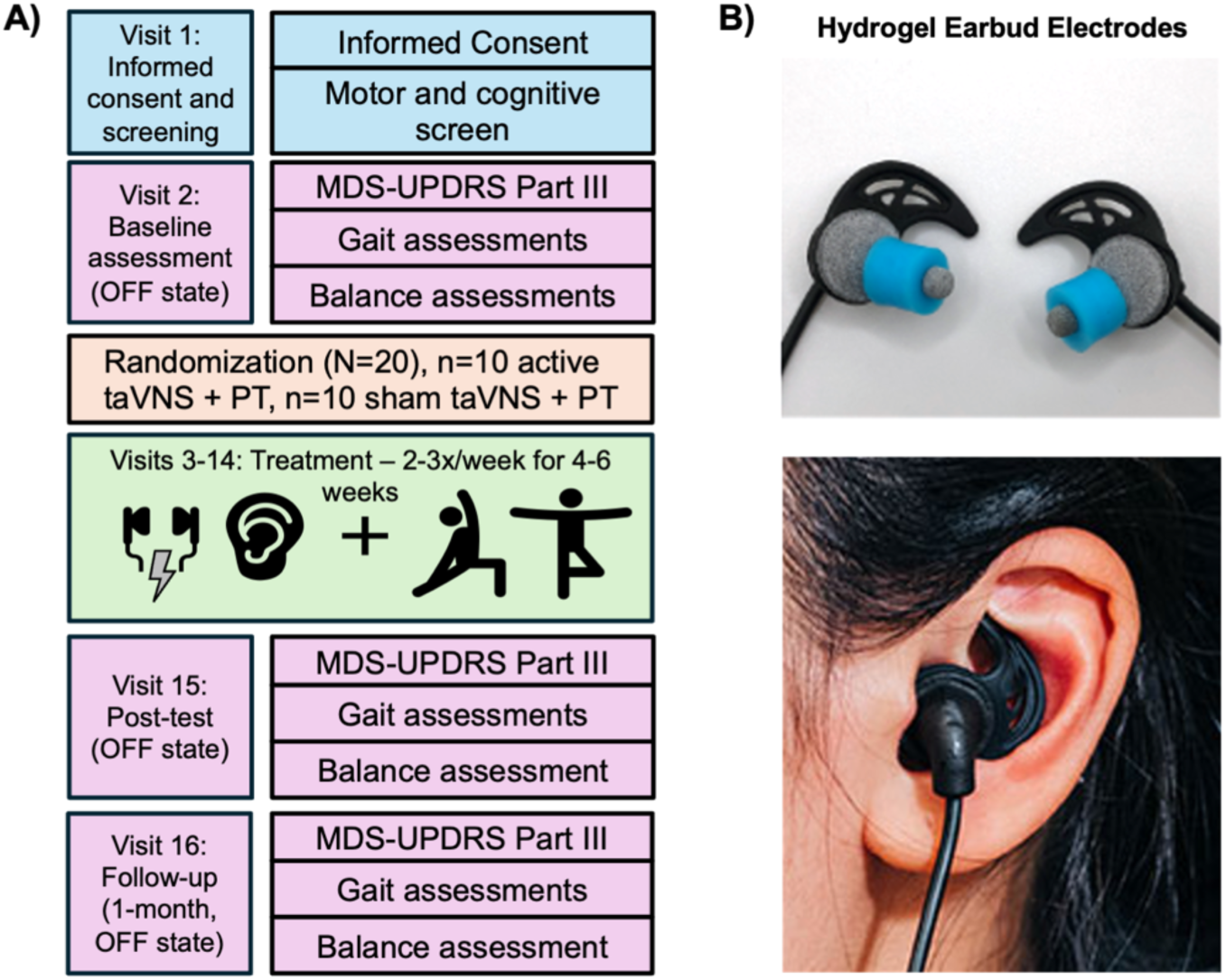
Overview of Study Design and Methods. **A)** The block diagram illustrates subject flow and treatment through the feasibility evaluation of bilateral transcutaneous auricular vagus nerve stimulation combined with physical therapy for improving Parkinsonian symptoms. **B)** Hydrogel earbud electrodes used to in this study that were designed to bilaterally target cranial nerve afferents innervating the skin of the external acoustic meatus.

### 2.2 Participants

Participants were recruited through UAB’s Movement Disorders Clinic, local support groups, and patient advocacy organizations. Inclusion criteria were (1) neurologist-confirmed diagnosis of idiopathic Parkinson’s disease, (2) age 35–80, (3) stable medications, (4) no falls in 6 weeks, (5) ability to walk without aids, and (6) written approval from the treating neurologist to participate in the study. Exclusion criteria were: (1) moderate-severe cognitive impairment (MoCA <20); (2) psychiatric comorbidities; (3) clinically significant cardiovascular or neurological comorbidities (e.g., history of myocardial infarction, uncontrolled hypertension, heart failure, significant arrhythmia, stroke, or transient ischemic attack), as determined by chart review and patient history; (4) implanted medical devices; (5) history of seizures and/or vasovagal syncope; and (6) pregnancy. Interested participants first completed a phone eligibility screen. Those eligible were invited for in-person screening and consent. After eligibility was confirmed and physician approval obtained, participants were consented and enrolled (**Figure 1**).

### 2.3 Interventions

Participants completed 12 sessions (2–3/week) over six weeks reflecting typical PT care. Each 60-minute session began with baseline heart rate (HR) and blood pressure (BP) reading, followed by 15 minutes of active or sham taVNS while resting. Study staff remained present and reassessed HR/BP post-stimulation. Participants then completed a 45-minute one-on-one PT session (see below, PWR! Moves PT Treatment) with a PD-trained clinician, followed by post-exercise vitals (**Figure 2A**). The only difference between groups was the electrical current output of the taVNS device.

#### 2.3.1 Transcutaneous Auricular Vagus Nerve Stimulation (taVNS)

Bilateral taVNS was delivered using a small, current-controlled device (vagus.net) connected to conductive, hydrogel earbud electrodes (BRAIN Buds; IST, LLC) inserted into the external acoustic meatus of the right and left ears (**Figure 2B**). Skin was inspected before and after each use. Biphasic pulses (250 µsec pulse durations, 50 µsec inter-pulse interval) were delivered at 30 Hz and 1.0 to 4.0 mA for 15 minutes. This stimulation duration was selected to balance feasibility within a 60-minute PT session and to align with prior clinical and preclinical taVNS studies in PD and other neurological populations, which have demonstrated acute physiological effects at similar durations [37, 39, 41, 48, 49]. Stimulation intensity was individually titrated for comfort at each session, beginning at 4.0 mA. Most participants found this intensity tolerable, however if discomfort was reported, stimulation intensity was gradually decreased in 0.1–0.5 mA increments until comfort was achieved. Our rationale for this comfort threshold approach was to ensure tolerability across repeated sessions in a pragmatic, clinic-based context. Sham devices mimicked controls but delivered 0 mA current.

#### 2.3.2 PWR! Moves Physical Therapy Treatment

PWR! Moves^®^ (pwr4life.org) is a flexible and adaptable PD-specific exercise program that is increasingly utilized in PT practice. This program was selected as the standard exercise platform for this study to provide a relevant foundation for testing whether adjunctive taVNS can enhance motor outcomes in the clinical setting. Participants in both groups completed 45-minute PWR! Moves PT session with a PWR! Moves Certified Clinician. The program includes 4 basic movements labeled PWR! Up, Rock, Twist, and Step, targeting antigravity extension, weight shifting, axial mobility, and transitional movements. Each movement can be performed in standing, sitting, quadruped, prone, or supine positions. Sessions were tailored to each participant’s ability level, with emphasis on large amplitude and high effort during sessions. Progression and prescription of all movements were determined by the clinician. Patient specific deficits (whether self-reported or noted during motor function assessments) were considered in treatment sessions.

### 2.4 Outcome Measures

Safety and feasibility metrics included 1) recruitment, enrollment, and attrition rates; 2) adverse events experienced during stimulation or exercise including autonomic symptom monitoring; and 3) treatment tolerability [50]. Changes in the Movement Disorders Society Unified Parkinson’s Disease Rating Scale Motor Sub-Scale (MDS-UPDRS Part III) were assessed over three time points to assess trends in treatment response: 1) baseline (assessment 1), 2) immediately following 12 taVNS + PT sessions (assessment 2), and at 4-weeks follow-up (assessment 3). The MDS-UPDRS Part III was administered at the start of each visit to minimize fatigue effects. Additional outcome measures included a series of clinical tests commonly used in PT practice, including the Modified Clinical Test of Sensory Interaction and Balance (mCTSIB; measure of static balance), Functional Gait Assessment (FGA; measure of dynamic balance, score out of 30, higher scores indicate better balance), Mini Balance Evaluation Systems Test (mini-BEST; measure of static and dynamic balance, score out of 28, higher scores indicate better balance), Six-Minute Walk Test (6MWT; measure of CV endurance, measures distance walked in 6 minutes in feet), and the 10-meter walk test (10MWT-ss and 10MWT-fs, which measures the time taken to walk 10 meters at self-selected and fastest gait speeds). Additionally, CV response to stimulation and exercise was measured at each treatment visit by recording changes in HR, systolic blood pressure (SBP), and diastolic blood pressure (DBP) at three time points: 1) pre-taVNS, 2) immediately post-taVNS, and 3) immediately post-exercise. All data was collected on-site at UAB’s Wellness, Health, and Research Facility (WHARF).

### 2.5 Study Participants

We targeted a sample size of 20 (10 per group) based on the projected number of individuals that could be recruited, screened, enrolled, and retained within the study period. The study PI enrolled participants and assigned participants to interventions using RedCap’s randomization module. Participant blinding was attempted by using a standardized blinding script to minimize participant awareness of group assignment. Due to the small study team and limited personnel resources, the PI was not blinded to group assignment. All other study staff involved in assessments and intervention delivery remained blinded throughout the study to reduce potential bias.

### 2.6 Statistical Analysis

Feasibility and safety outcomes including recruitment, enrollment, attrition, and adverse events were summarized using frequency counts and percentages. Descriptive statistics were calculated for baseline demographic and clinical variables. Baseline differences and tolerability were compared using independent t-tests and chi-square tests, as appropriate.

This study was not statistically powered to evaluate efficacy; however, we conducted ancillary and exploratory analyses to identify potential trends in motor and CV outcomes and to gather preliminary effect size estimates to guide protocol refinement and outcome selection for a future, adequately powered clinical trial. Within-group analyses of motor outcomes (MDS-UPDRS III, gait, and balance) are reported descriptively and included repeated measures ANOVAs with Bonferroni corrections for multiple comparisons as appropriate. CV responses (HR, SBP, DBP) were analyzed using paired t-tests for pre/post-taVNS and pre-taVNS/post-exercise intervals. Effect sizes were reported using Cohn’s d along with 95% confidence intervals. Between-group comparisons of change scores were conducted for outcomes showing within-group significance using two-tailed t-tests. Change scores for motor outcomes were calculated from baseline to post-treatment (immediate effect) and from baseline to follow-up (sustained effect). Change scores for CV outcomes were calculated from pre-taVNS to post-taVNS (to elucidate effect of taVNS alone) and from pre-taVNS to post-exercise (to elucidate effect of taVNS plus PT). Analyses were conducted using JASP (version 0.18.3), with significance set at p < 0.05.

## 3. RESULTS

### 3.1 Feasibility

We screened 44 individuals of which 33 met preliminary criteria and 26 were consented and enrolled between May 2023 and Sept 2024. The study was completed once recruitment goals were met. Three subjects were excluded after in-person screening, yielding a 70% enrollment rate (23/33). One participant was excluded pre-randomization due to hypertension. Twenty-two participants were randomized (11 per group). One sham participant withdrew due to an unrelated illness, and one active participant missed follow-up, resulting in a 91% completion rate and 95% adherence. A total of 20 participants (10 per group) were included in the final analyses. See **Figure 1** for the CONSORT diagram.

### 3.2 Demographics and Clinical Characteristics

Baseline demographic and clinical characteristics are presented in **Table 1**. Participants averaged 67.2 ± 7.1 years of age, with PD duration of 5.0 ± 3.2 years and mild-moderate disease severity (H&Y 1-3). There were no significant between-group differences in age (p=0.143, CI: −1.75, 11.15), years since diagnosis (p=0.813, CI: −3.42, 2.72), H&Y stage (p=0.382, CI: −3.42, 2.72), cognitive function (MoCA OFF: p=0.640, CI: −2.71, 1.71; MoCA ON: p=0.723, CI: −2.74, 1.94), and ethnic composition (p=0.305). Sex and gender distributions were identical across groups. There were no significant differences in MDS-UPDRS Part III scores at baseline in the OFF- (sham: 36.7 ± 8.6, active: 31.0 ± 12.68) and ON-medication (sham: 34.2 ± 14.66, active: 26.3 ± 13.21) states (OFF: p=0.255, CI: −4.48, 15.88; ON: p=0.222, CI: −5.21, 21.01).

**Table 1.** Demographics and Clinical Characteristics by Treatment Group.

| Characteristic | Total (N=20) | Sham (n=10) | Active (n=10) | p-value | 95% CI |
| --- | --- | --- | --- | --- | --- |
| Age, years | 67.2 (7.10) | 69.5 (6.15) | 64.8 (7.51) | 0.143 | -1.75, 11.15 |
| Years since diagnosis | 5.0 (3.18) | 4.8 (3.05) | 5.2 (3.47) | 0.813 | -3.42, 2.72 |
| Hoehn & Yahr stage | 2.7 (0.75) | 2.8 (0.63) | 2.5 (0.85) | 0.382 | -0.40, 1.00 |
| MDS-UPDRS Part III (Off) | 33.9 (10.95) | 36.7 (8.6) | 31.0 (12.68) | 0.255 | -4.48, 15.88 |
| MDS-UPDRS Part III (On) | 30.3 (14.17) | 34.2 (14.66) | 26.3 (13.21) | 0.222 | -5.21, 21.01 |
| MoCA (Off) | 27.0 (2.31) | 26.7 (1.95) | 27.2 (2.70) | 0.640 | -2.71, 1.71 |
| MoCA (On) | 26.7 (2.43) | 26.5 (2.37) | 26.9 (2.60) | 0.723 | -2.74, 1.94 |
| Sex |  |  |  |  |  |
| Male | 16 (80.0%) | 8 (80.0%) | 8 (80.0%) | 1.000 |  |
| Female | 4 (20.0%) | 2 (20.0%) | 2 (20.0%) |  |  |
| Gender |  |  |  |  |  |
| Male | 16 (80.0%) | 8 (80.0%) | 8 (80.0%) | 1.000 |  |
| Female | 4 (20.0%) | 2 (20.0%) | 2 (20.0%) |  |  |
| Ethnicity |  |  |  |  |  |
| White/Caucasian | 19 (95.0%) | 10 (100.0%) | 9 (90.0%) | 0.305 |  |
| Black/African-American | 1 (5.0%) | 0 (0.0%) | 1 (10.0%) |  |  |

### 3.3 Safety and Tolerability of Treatment

There were no adverse events reported throughout the duration of the study that were directly attributable to the study treatment. Tolerability results (**Table 2**) showed participants in the active group were more likely to perceive the stimulation (p=0.021). We found 33.3% of participants in the active group reported feeling the stimulation every time and 56.6% most of the time, while the sham group reported only occasional (50.0%) or rare (50.0%) perception of stimulation. Perceived stimulation intensity ratings (0–10 scale) were higher in the active group (3.0 ± 1.89) compared to the sham group (0.9 ± 1.29; p=0.009, CI: −3.62, −0.58), however an average rating of 3.0 in the active group indicates that the intensity was perceived as relatively mild. Tingling was the most frequently reported sensation (70% active vs. 40% sham), followed by a pulsating sensation (30% active, 0% sham). No participants reported pain or discomfort. As these results indicate, blinding integrity was not maintained, with participants in the active group more likely to correctly identify their stimulation status.

**Table 2.** Tolerability of Bilateral Transcutaneous Auricular Vagus Nerve Stimulation.

|  | **Sham (n=10)** | **Active (n=10)** | **p-value** |
| --- | --- | --- | --- |
| **Feels Stimulation** |  |  | **0.021\*** |
| Yes | 0 (0.0%) | 4 (40.0%) |  |
| Sometimes | 4 (40.0%) | 4 (50.0%) |  |
| No | 6 (60.0%) | 1 (10.0%) |  |
| **If yes/sometimes, frequency felt** |  |  | **0.020\*** |
| Every Time | 0 (0.0%) | 3 (33.3%) |  |
| Most of the Time | 0 (0.0%) | 5 (55.6%) |  |
| Occasionally | 2 (50.0%) | 1 (11.1%) |  |
| Rarely | 2 (50.0%) | 0 (0.0%) |  |
| **Stimulation Intensity (0–10)** | 0.9 (1.29) | 3.0 (1.89) | **0.009\*** |
| **Tingling Sensation** |  |  | 0.178 |
| Yes | 4 (40.0%) | 7 (70.0%) |  |
| No | 6 (60.0%) | 3 (30.0%) |  |
| **Pulsating Sensation** |  |  | 0.060 |
| Yes | 0 (0.0%) | 3 (30.0%) |  |
| No | 10 (100.0%) | 7 (70.0%) |  |
| **Warm Sensation** |  |  | 1.000 |
| Yes | 0 (0.0%) | 0 (0.0%) |  |
| No | 10 (100.0%) | 10 (100.0%) |  |
| **Pressure** |  |  | 1.000 |
| Yes | 0 (0.0%) | 0 (0.0%) |  |
| No | 10 (100.0%) | 10 (100.0%) |  |
| **Pain** |  |  | 1.000 |
| Yes | 0 (0.0%) | 0 (0.0%) |  |
| No | 10 (100.0%) | 10 (100.0%) |  |
| **Dull Ache** |  |  | 1.000 |
| Yes | 0 (0.0%) | 0 (0.0%) |  |
| No | 10 (100.0%) | 10 (100.0%) |  |
| **Numbness** |  |  | 1.000 |
| Yes | 0 (0.0%) | 0 (0.0%) |  |
| No | 10 (100.0%) | 10 (100.0%) |  |
| **Overall Tolerability of Treatment** |  |  | 0.060 |
| Very Comfortable | 8 (80.0%) | 6 (60.0%) |  |
| Comfortable | 0 (0.0%) | 3 (30.0%) |  |
| Neutral | 2 (20.0%) | 1 (10.0%) |  |
| Uncomfortable | 0 (0.0%) | 0 (0.0%) |  |
| Very Uncomfortable | 0 (0.0%) | 0 (0.0%) |  |

### 3.4 Effects of taVNS combined with Exercise on Motor Function

We were most interested in how treatment affected individual changes from their own baseline, so first analyzed within-group outcomes. Repeated measures ANOVAs revealed that both sham and active taVNS groups experienced improvement in MDS-UPDRS Part III scores over time (active taVNS: p = 0.011, ηp² = 0.395; sham taVNS: p = 0.016, η_p_² = 0.370, **Figure 3A**). Post-hoc analyses revealed a significant reduction from baseline to post-treatment in both groups (active: p = 0.034; sham: p = 0.014), likely reflecting the short-term benefits of PT. However, only the active taVNS group maintained statistically significant improvements at the one-month follow-up (baseline vs. follow-up: p = 0.018), whereas the sham group exhibited a regression toward baseline motor function (p = 0.500; **Figure 3A**). In the sham group, there were no other significant changes in motor function over time (**Table 3**).

**Figure 3.**
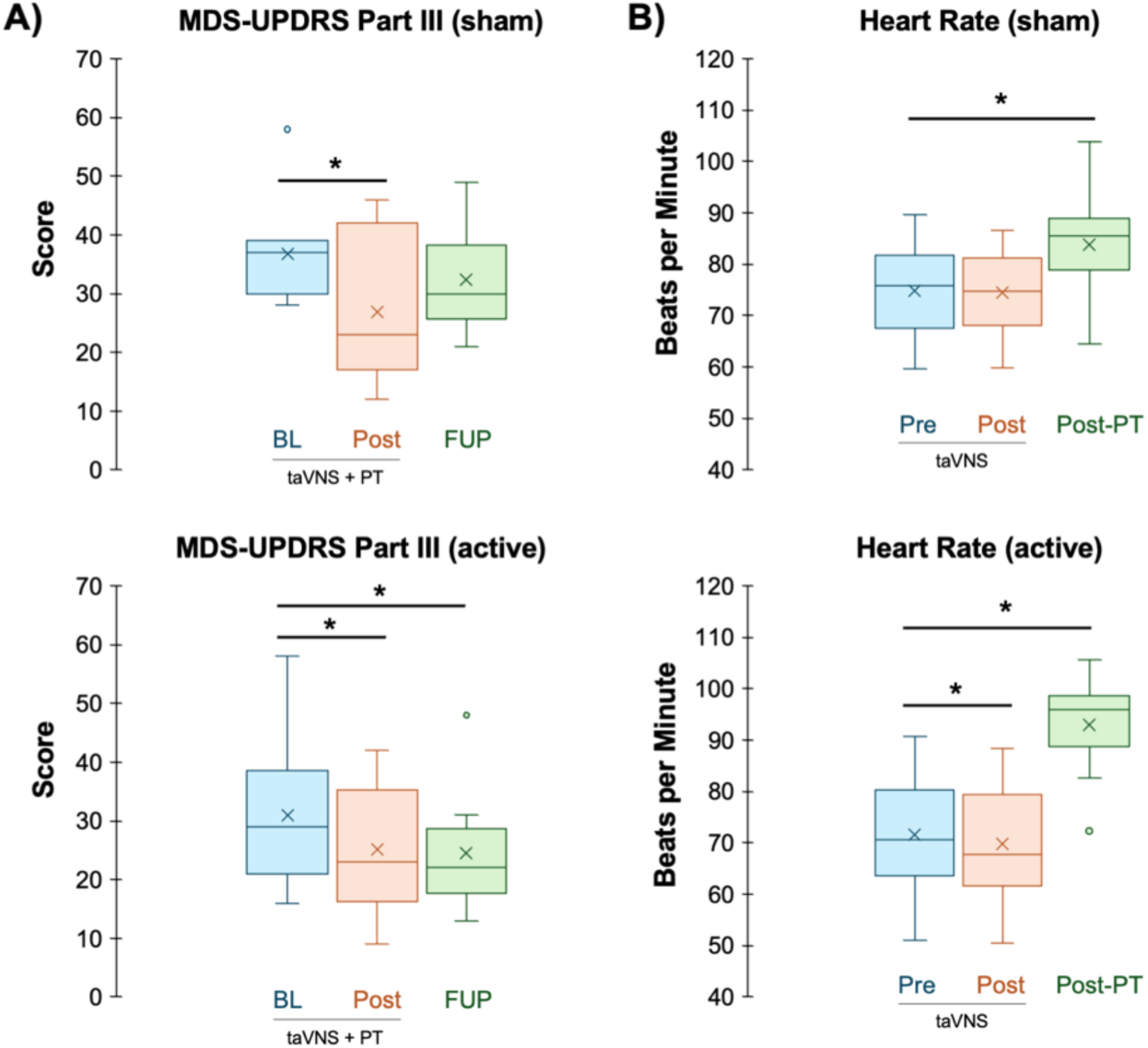
Influence of Treatment on Motor Symptoms and Heart Rate. **A**) The histograms illustrate MDS-UPDRS Part III scores across time. Scores were assessed at baseline (*BL*), following 12 transcutaneous auricular vagus nerve stimulation (taVNS) plus physical therapy treatment (PT) exercise sessions over six weeks (*Post)*, and one-month follow-up (*FUP*). Within-group repeated-measures ANOVA revealed a significant main effect of time for both groups (active taVNS: F(2,18) = 5.89, p = 0.011; sham taVNS: F(2,18) = 5.28, p = 0.016). Post hoc comparisons showed significant improvements from baseline to post-test in both groups (active: p = 0.034; sham: p = 0.014), but only the active taVNS treatment group maintained significant improvements follow-up (p = 0.018). **B)** Average heart rate (HR) during treatment sessions. HR was measured at each visit before (*Pre*) and after (*Post*) taVNS treatment just prior to exercise, as well as immediately post-exercise (*Post-PT*) and averaged across 12 sessions for each participant. The active taVNS group, but not the sham group, exhibited a significant decrease in HR in response to taVNS treatment prior to exercise. Paired-sample t-tests also revealed significant increases in HR from pre-taVNS to post-exercise in both groups (active taVNS: p < 0.001, d = –2.87; sham taVNS: p < 0.001, d = –1.56), consistent with the effects of exercise. An *asterisk* indicates p < 0.05.

**Table 3.**
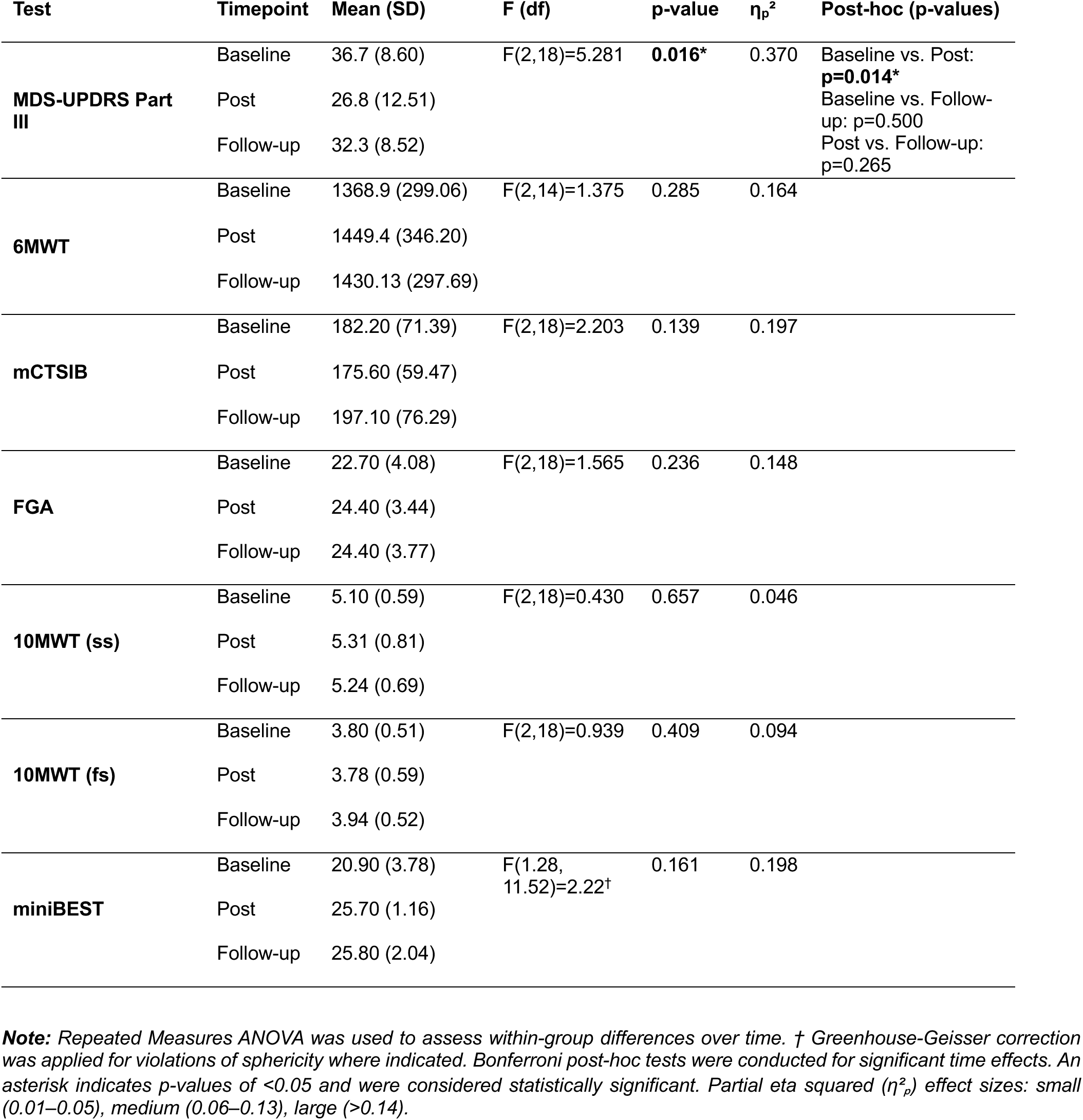
Motor Outcomes in Response to Sham Treatment.

In contrast, the active taVNS group exhibited a significant increase in distance walked during the 6MWT (baseline: 1390.04 ± 126.22, post-treatment: 1556.00 ± 196.41, follow-up: 1505.22 ± 173.10; p = 0.045, η_p_² = 0.386; **Table 4**). Post-hoc comparisons show a significant improvement from baseline and post-treatment (p = 0.020). However, this improvement was not sustained at follow-up (p = 0.173), which may suggest that any potential benefits to CV endurance were short-lived in the absence of continued intervention. Significant improvements in dynamic balance were also observed, as measured by the FGA (baseline: 25.00 ± 3.43, post-test: 28.60 ± 2.17, follow-up: 27.69 ± 2.80; p = 0.001, η_p_² = 0.531) and the Mini-BESTest (baseline: 22.30 ± 2.54, post-test: 25.70 ± 1.16, follow-up: 25.80 ± 2.04, p < 0.001, η_p_² = 0.640). Post-hoc analyses revealed immediate improvements from baseline to post-test (FGA: p = 0.001; Mini-BESTest: p < 0.001), and importantly, these gains were sustained at follow-up (FGA: p = 0.016; Mini-BESTest: p < 0.001), suggesting potentially enduring changes in balance performance (**Table 4**). Likely due to the small sample size used in this feasibility trial, we observed no significant differences between treatment groups for the MDS-UPDRS Part III, 6MWT, FGA, or mini-BEST test (**Table 6**).

**Table 4.**
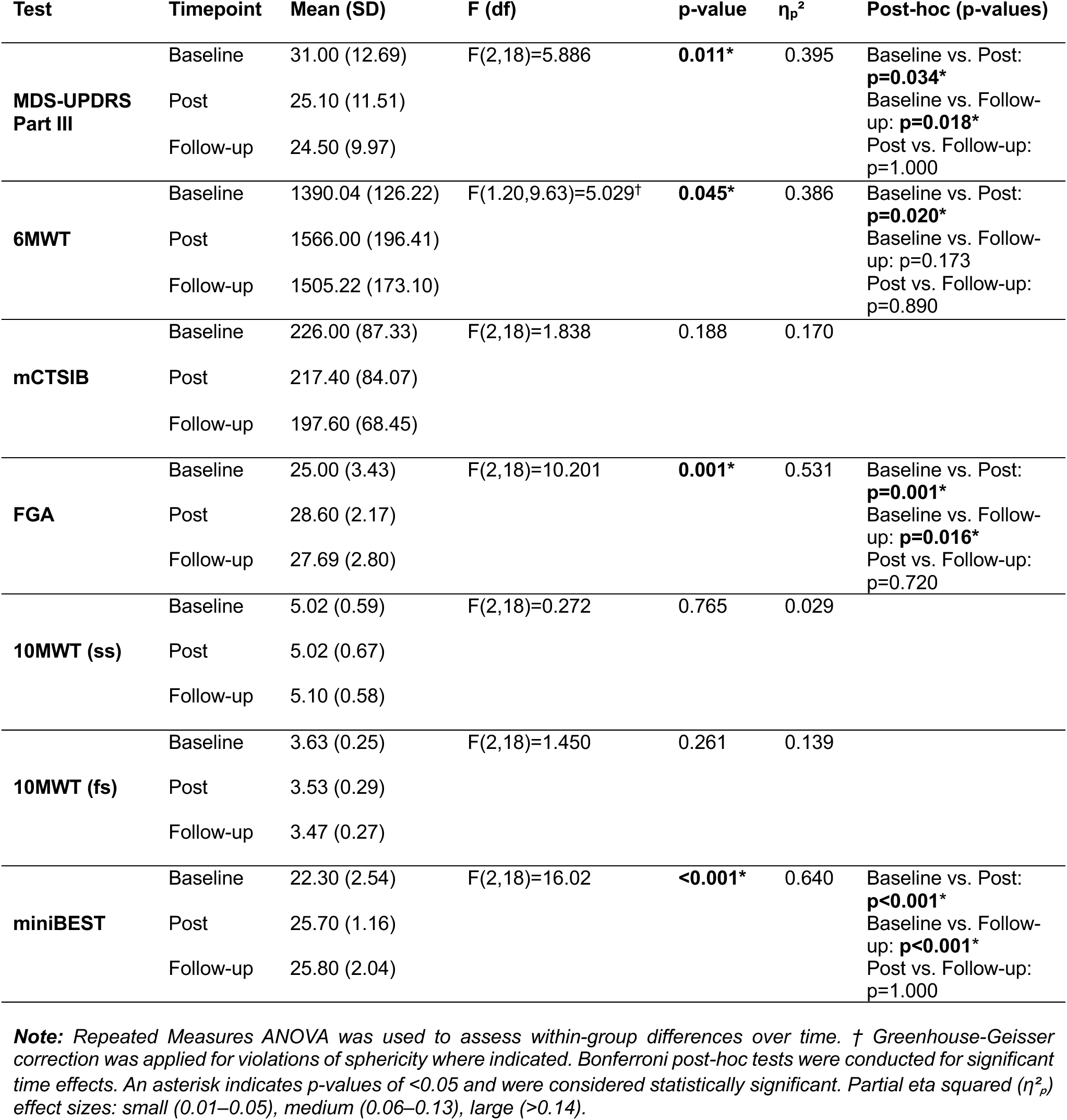
Motor Outcomes in Response to Active Treatment.

### 3.5 Effects of taVNS and Exercise on Cardiovascular Activity

A summary of CV activity including HR, DBP, and SBP changes in response to taVNS and exercise for both treatment groups is presented in **Table 5**. In the sham group, only SBP decreased significantly post-taVNS (pre-sham SBP: 128.53 ± 12.50, post-sham: 120.84 ± 11.78; p=0.023, CI: 2.17, 13.21, d = 0.996). In contrast, active taVNS significantly reduced SBP, DBP, and HR, all with large effect sizes (pre-active SBP: 130.21 ± 9.86, post-active SBP: 121.50 ± 6.93, p<0.001, CI: 5.11, 12.31, d: 1.729; pre-active DBP: 75.93 ± 6.07, post-active DBP: 73.09 ± 5.04, p=0.003, CI: 1.26, 4.42, d: 1.284; pre-active HR: 71.54 ± 12.25, post-active HR: 69.85 ± 11.44, p=0.027, CI: 0.24, 3.15, d: −0.834; **Table 5**).

**Table 5.** Effects on Cardiovascular Outcomes Within Treatment Groups.

| Condition | Mean (SD) | p | 95% CI | Cohn's d |
| --- | --- | --- | --- | --- |
| **Sham taVNS Group** |  |  |  |  |
| **SBP (mmHg)** |  |  |  |  |
| Pre taVNS | 128.53 (12.50) | **0.012\*** | 2.17, 13.21 | 0.996 |
| Post taVNS | 120.84 (11.78) |  |  |  |
| Post exercise | 134.01 (14.91) | 0.058 | -11.18, 0.23 | -0.687 |
| **DBP (mmHg)** |  |  |  |  |
| Pre taVNS | 76.76 (6.19) | 0.255 | -1.13, 3.76 | 0.385 |
| Post taVNS | 75.44 (6.54) |  |  |  |
| Post exercise | 78.05 (7.20) | 0.099 | -2.88, 0.30 | -0.582 |
| **HR (BPM)** |  |  |  |  |
| Pre taVNS | 74.91 (9.61) | 0.367 | -0.66, 1.61 | 0.300 |
| Post taVNS | 74.44 (8.96) |  |  |  |
| Post exercise | 83.89 (11.29) | **<0.001\*** | -13.09, -4.86 | -1.559 |
| **Active taVNS Group** |  |  |  |  |
| **SBP (mmHg)** |  |  |  |  |
| Pre taVNS | 130.21 (9.86) | **<0.001\*** | 5.11, 12.31 | 1.729 |
| Post taVNS | 121.50 (6.93) |  |  |  |
| Post exercise | 137.60 (15.11) | 0.056* | -15.02, 0.24 | -0.693 |
| **DBP (mmHg)** |  |  |  |  |
| Pre taVNS | 75.93 (6.07) | **0.003\*** | 1.26, 4.42 | 1.284 |
| Post taVNS | 73.09 (5.04) |  |  |  |
| Post exercise | 77.26 (6.68) | 0.235 | -3.69, 1.03 | -0.403 |
| **HR (BPM)** |  |  |  |  |
| Pre taVNS | 71.54 (12.25) | **0.027\*** | 0.24, 3.15 | 0.834 |
| Post taVNS | 69.85 (11.44) |  |  |  |
| Post exercise | 92.91 (9.42) | **<0.001\*** | -26.71, -16.04 | -2.865 |

**Note:** Paired sample *t*-tests were used to assess changes in SBP, DBP, and HR over time within each treatment group. The first comparison assessed changes in SBP, DBP and HR from pre-taVNS to post-taVNS. The second comparison assessed changes in SBP, DBP, and HR from pre-taVNS to post-exercise. Effect sizes were calculated using Cohn's *d*, where values of 0.1–0.3 indicate a small effect, 0.3–0.5 indicate a medium effect, and greater than or equal to 0.5 indicate a large effect. An asterisk indicates a *p*-value of less than 0.05 and was considered statistically significant.

As expected, both groups showed HR increases immediately following exercise (sham: p<0.001, CI: −13.09, −4.86, d = −1.559; active: p<0.001, CI: −26.71, −16.04, d = −2.865, **Figure 3B**). However, between-group comparisons indicate that this difference was statistically significant between groups (p<0.001, CI: −18.66, −6.14, d: −1.86, **Table 6**), with the active taVNS group demonstrating a more robust increase in HR (21.37 ± 7.46 BPM) in response to exercise compared to sham treatment changes in HR (8.98 ± 5.76 BPM). These findings suggest that taVNS may influence CV function at rest and may result in a more pronounced HR response to exercise when administered immediately beforehand (**Figure 3B**), however these results should be interpreted cautiously, and larger confirmatory trials are needed to verify this observation.

**Table 6.** Effects on Motor and Cardiovascular Outcomes Across Treatment Conditions.

| Test | Timepoint | Mean (SD) | p-value | 95% CI | Cohn's d |
| --- | --- | --- | --- | --- | --- |
| **MDS-UPDRS Part III** | Δ Baseline to Post | Sham: -9.90 (8.63)<br>Active: -5.90 (8.03) | 0.298 | -11.84, 3.84 | -0.480 |
|  | Δ Baseline to F/U | Sham: -4.40 (8.67)<br>Active: -6.50 (5.67) | 0.529 | -4.78, 8.98 | 0.287 |
| **6MWT** | Δ Baseline to Post | Sham: 64.67 (162.94)<br>Active: 175.96 (159.01) | 0.162 | -272.17, 49.59 | -0.691 |
|  | Δ Baseline to F/U | Sham: 83.89 (150.22)<br>Active: 132.06 (109.55) | 0.432 | -174.46, 78.12 | -0.370 |
| **FGA** | Δ Baseline to Post | Sham: 1.70 (3.43)<br>Active: 3.60 (2.32) | 0.164 | -4.65, 0.85 | -0.649 |
|  | Δ Baseline to F/U | Sham: 1.30 (3.27)<br>Active: 2.60 (3.17) | 0.378 | -4.32, 1.72 | -0.404 |
| **Mini-BEST** | Δ Baseline to Post | Sham: 1.90 (3.57)<br>Active: 3.40 (2.46) | 0.289 | -4.38, 1.38 | -0.489 |
|  | Δ Baseline to F/U | Sham: 1.30 (3.23)<br>Active: 2.60 (3.17) | 0.106 | -4.92, 0.52 | -0.761 |
| **SBP (mmHG)** | Δ pre-taVNS to post-taVNS | Sham: -7.69 (7.72)<br>Active: -8.71 (5.04) | 0.730 | -5.10, 7.15 | 0.157 |
|  | Δ pre-taVNS to post-exercise | Sham: 5.48 (7.97)<br>Active: 7.39 (10.67) | 0.655 | -10.76, 6.94 | -0.203 |
| **DBP (mmHG)** | Δ pre-taVNS to post-taVNS | Sham: -1.32 (3.42)<br>Active: -2.84 (2.21) | 0.252 | -1.18, 4.23 | 0.529 |
|  | Δ pre-taVNS to post-exercise | Sham: 1.29 (2.22)<br>Active: 1.33 (3.30) | 0.987 | -2.68, 2.61 | -0.013 |
| **HR (BPM)** | Δ pre-taVNS to post-taVNS | Sham: -0.48 (1.58)<br>Active: -1.70 (2.03) | 0.151 | -0.49, 2.93 | 0.671 |
|  | Δ pre-taVNS to post-exercise | Sham: 8.98 (5.76)<br>Active: 21.37 (7.46) | **<0.001\*** | -18.66, -6.14 | -1.860 |

**Note:** Independent samples *t*-tests were used to assess between-group differences. For all motor outcomes (MDS-UPDRS Part III, 6MWT, FGA, and miniBEST), the first comparison evaluates differences in change scores from baseline to post-test assessments. The second comparison evaluates differences in change scores from baseline to follow-up. For all cardiovascular outcomes (SBP, DBP, and HR), the first comparison evaluates differences in changes scores from pre-taVNS to post-taVNS. The second comparison evaluates differences in change scores from pre-taVNS to post-exercise. Effect sizes were calculated using Cohn's, where values of 0.1–0.3 indicate a small effect, 0.3–0.5 indicate a medium effect, and greater than or equal to 0.5 indicate a large effect. An asterisk indicates a *p*-value of less than 0.05 and was considered statistically significant.

## 4. DISCUSSION

### 4.1 Overview

PT is a widely prescribed, non-pharmacologic therapy for PD. Exercise promotes increased neurotrophic factor expression [51], dopamine transmission [52], corticomotor excitability [53], and grey matter volume [54] in PD patients. However, the exercise intensity and frequency that drives these physiologic changes can be difficult to achieve in real-world clinical settings because of interference from time constraints, burden of care, and dysautonomia or impaired cardiovascular dynamics. Therefore, interventions that address these barriers could increase the translation of exercise research into clinical practice. Transcutaneous auricular Vagus Nerve Stimulation (taVNS) is a non-invasive neuromodulation method that could bridge this gap. This pilot study assessed the safety and feasibility of a combined taVNS plus PT intervention and identified trends in outcomes to inform future study design.

#### 4.1.1 Feasibility, Safety & Tolerability of Combining Bilateral taVNS with Exercise in PD

This pilot trial demonstrates that combining bilateral taVNS with PT is feasible, safe, and well-tolerated in individuals with mild-moderate PD. This observation is consistent with current literature supporting the safety of taVNS in both healthy adults [12] and individuals with PD [37, 38, 55]. Our study was the first to formally evaluate safety and tolerability of combining taVNS with exercise in PD. High recruitment, enrollment and adherence rates were achieved, and zero adverse events occurred that were attributable to the taVNS combined with PT treatment. The results of the tolerability survey suggest that our taVNS approach is comfortable and acceptable from a patient experience standpoint, which is critical for clinical translation, as interventions that are not well-tolerated or perceived positively by patients are unlikely to be adopted. These findings justify a larger clinical trial to formally assess efficacy of this intervention.

#### 4.1.2 Influence of taVNS on Parkinsonian Symptoms

Although this pilot study was not powered to detect definitive between-group differences, exploratory analyses revealed patterns consistent with our hypothesis that taVNS may augment PT outcomes in PD. Both active and sham taVNS combined with PT improved MDS-UPDRS Part III scores immediately post-intervention, reflecting expected benefits of rehabilitation that are well-documented in PD [56]; however, only the active group maintained these gains at the 4-week follow-up, suggesting the possibility of increased retention of motor improvements when taVNS is added. Improvements in dynamic balance (FGA and Mini-BEST) were also exclusive to the active taVNS treatment group and persisted at follow-up. This may reflect a synergistic interaction between the LC–norepinephrine (LC-NE) mediated neuroplastic effects of taVNS [42–45, 57] and the task-specific neuroplasticity induced by PWR! Moves, which emphasizes large-amplitude, dynamic balance training. One plausible mechanism to support these results is that taVNS primes the brain by activating the LC-NE system prior to therapy, increasing arousal, facilitating synaptic plasticity, and enhancing consolidation of the motor skills practiced during PT. This is a likely contributing mechanism given noradrenergic signaling is known to be disrupted in patients with PD [3, 36]. Another possibility parallels findings from stroke rehabilitation research, where VNS paired with task-specific training produced more robust and durable motor gains than training alone [58]. It is not known how the changes we observed in a priming approach (delivering taVNS prior to task engagement) may compare to the pairing of taVNS stimulation with successful movement execution in an event-based manner. Determining whether taVNS priming, pairing, or a combination of approaches yields the most potent neuroplastic response represents an important avenue for future research.

We also observed that taVNS was associated with acute modulation of cardiovascular (CV) autonomic function at rest, including reductions in SBP, DBP, and HR. Given the high prevalence of orthostatic hypotension and syncope in PD [59], the clinical desirability of these resting changes warrants consideration. However, in this study, the effects observed were modest, transient, and not associated with adverse events. Importantly, administering taVNS immediately before exercise elicited a more robust HR response during activity. Similar observations have been reported in response to bilateral taVNS before, during, and following exercise paradigms in healthy individuals [23, 24]. Hatik and colleagues (2023) first reported that bilateral taVNS was significantly more effective than left unilateral taVNS at reducing fatigue and pain during exercise while significantly decreasing HR, SBP, and DBP across treatment days [24]. More recently, Ackland and colleagues (2025) reported that bilateral taVNS can lead to significantly reduced fatigue, increased work rate, and enhanced CV dynamics including increased HR and VO_2_ max during exercise compared to controls [23]. These observations are consistent with known mechanistic roles of cardiac vagal activity in exercise performance [17, 27]. Thus, the more dynamic CV response we observed may reflect improved autonomic adaptability, which could enhance exercise tolerance and allow patients to engage more fully in therapy sessions, thereby indirectly contributing to motor improvements. This interpretation is further supported by the observation that only the active taVNS group demonstrated improvements in the 6MWT. However, these endurance gains were not maintained at the 4-week follow-up, suggesting that ongoing or repeated stimulation may be necessary to sustain CV benefits. This is also consistent with previous observations that the cardiorespiratory benefits produced by bilateral taVNS during exercise in healthy individuals do not have carryover effects and more likely reflect acute effects on CV function [23]. Considering that individuals with PD often exhibit a blunted HR response to exercise [60–62], our findings point to a potentially valuable role for taVNS in acutely modulating autonomic cardiac vagal function to optimize exercise capacity.

The preliminary effects we observed on HR and BP raise other mechanistic issues worth noting. First, we bilaterally targeted cranial nerve fibers innervating the external acoustic meatus (EAM) of subjects. This anatomical location of the ABVN and methods have been shown to modulate plasticity and CV activity by engaging NA signaling [13, 63–66]. This location is also partially innervated by the auriculotemporal branch (ATN) of the trigeminal nerve (cranial nerve V_III_) meaning the outcomes we observed with this embodiment of taVNS involve both trigeminal and vagal modulation [13]. Our observations of decreased HR in response to bilateral transcutaneous electrical nerve stimulation of the EAM are consistent with trigeminal cardiac and trigeminal vagal reflexes described throughout the literature, which have been shown to underlie the classic mammalian diving reflex [67–70]. Teasing apart the exact contributions of the various cranial nerves (V, VII, and X) and cervical nerves (great auricular nerve and lesser occipital nerve) affected by different external ear stimulation approaches is a hurdle for the neuromodulation field. It can be expected there is a large degree of individual variability, so it may be prudent to optimize human factors like comfort and ease of use which were key factors in our selection of skin-like hydrogel earbud electrodes designed specifically for bioelectronic medicine applications [13]. At a molecular level, the acute effects of taVNS to significantly reduce HR and BP resemble the actions of clonidine, an alpha-2 (α2) adrenergic receptor (AR) agonist. We have previously reported that trigeminal vagal stimulation can acutely reduce sympathetic tone while producing effects on psychophysiological arousal like clonidine [71]. This is more directly supported by observations in PET imaging showing that taVNS modulates NA (LC-NE) system activity, in part, by acting on α2-ARs [72]. More recently, it has been shown that α2-AR activation is necessary for the expression of taVNS induced motor plasticity [73]. Similarly, the β-AR receptor antagonist propranolol, which reduces sympathetic (NA) tone like the α2-AR agonist clonidine, has been reported to significantly enhance VNS-induced plasticity [74]. Collectively, these PET imaging and pharmacological data indicate that LC-NE modulation of AR activity is a primary mechanism by which taVNS bestows its effects on motor plasticity and possibly CV dynamics. Our exploratory findings are consistent with a dual-mechanism model in which taVNS may (1) engage neural circuitry that facilitates enhanced motor plasticity and (2) modulate CV autonomic function to support more effective participation in exercise. The transient nature of CV endurance improvements, contrasted with the sustained motor and balance gains, raises important questions about the relative contributions and interaction of these mechanisms. Future trials should incorporate pharmacology treatments with dynamic measures of autonomic function (e.g., HRV, VO₂ max) alongside direct markers of neuroplasticity to clarify the pathways by which taVNS influences rehabilitation outcomes in PD.

### 4.2 Limitations & Future Directions

This study has several potential limitations. First, the small sample size limits the statistical power and precludes definitive conclusions about efficacy. Second, although this was designed as a single-blinded pilot study, blinding integrity was difficult to maintain. Participants in the active group more frequently perceived stimulation compared to the sham group, indicating that the use of perceptible stimulation intensities may have compromised participant blinding and introduced expectancy or placebo effects. Consequently, protocol refinement is needed. Future studies should include separate, blinded assessors and consider sub-perceptual stimulation intensities or sham electrode placement at other external ear locations to ensure a truly double-blinded study design. Additionally, additional pulse profiles should be examined to identify optimal therapeutic dosage. Furthermore, only individuals with mild-moderate PD were enrolled, limiting generalizability to those with more advanced disease. The follow-up period was also relatively short, and longer-term follow-up outcomes (e.g., 6–12 months) are needed to assess the durability of treatment effects. Our observations with respect to CV function were assessed only at discrete time points (pre-taVNS, post-taVNS, and post-exercise). Continuous CV monitoring including heart rate variability or cardiometabolic testing may provide a more comprehensive understanding of how taVNS affects autonomic modulation. Additionally, only one exercise modality was used. While functionally oriented, it remains unclear whether other exercise modalities, such as CV endurance training or task-specific rehabilitation, may yield greater benefits when combined with taVNS. Finally, no direct measures of neuroplasticity were collected. Future trials incorporating neuroimaging or neurophysiological outcomes are needed to elucidate underlying mechanisms.

## 4.3 Conclusions

Combining taVNS with PT is feasible, safe, and tolerable for individuals with mild-moderate idiopathic PD. Exploratory findings also suggest that taVNS plus PT may be associated with trends toward greater and more sustained improvements in motor function compared to PT alone; however, the study was not powered to confirm these differences. Further clinical studies are required to draw definitive conclusions about efficacy. The results of this study provide preliminary evidence that taVNS combined with exercise is a is a feasible and potentially scalable therapeutic option for patients suffering from PD.

## Data Availability

All data produced in the present study are available upon reasonable request to the authors

## Acknowledgements

Funding was provided by the UAB School of Health Professions Junior Faculty Development Grant. This work has not been previously presented at a scientific meeting.

## Declaration of generative AI usage

The authors did not use generative AI in design or conceptualization of this research. The authors did not use generative AI to create or produce the ideas and content of this manuscript. During the preparation of this work the authors used OpenAI to check spelling and grammar. After using this tool/service, the authors reviewed and edited the content as needed and take full responsibility for the content of the published article.

